# Understanding healthcare workers’ experiences of face mask and RPE use in healthcare settings: an interview study

**DOI:** 10.1101/2023.11.20.23298509

**Authors:** Holly Carter, Ashley Sharp, Louise Davidson, Clare Foster, Emma McGuire, Colin Brown, Dale Weston

## Abstract

Whilst healthcare workers (HCWs) are at high risk of contracting COVID-19, measures can be put in place to reduce the spread of COVID-19 and other respiratory infections in healthcare settings. These currently include the use of masks: fluid-resistant surgical masks and respiratory protective equipment. However, for mask policies to be effective, compliance with their use must be high. This study interviewed 12 HCWs from a variety of backgrounds to understand their experiences of mask use. We explored factors associated with compliance with mask use and potential impacts on HCW wellbeing. Overall, participants reported good understanding of the benefits of masks and high compliance levels with policy. However, factors that reduced their compliance with mask policy and impacted their ability to carry out their role were highlighted. These included wearing masks for longer durations, policy being perceived as out of proportion with risk, communication challenges, and discomfort. This study highlights the importance of clear communication of guidance, particularly when it has changed, ensuring staff are familiar with up-to-date research on efficacy of masks, and ensuring guidance aligns with risk. Furthermore, this study highlights the importance of masks being required for an appropriate duration (based on risk).

## Introduction

There is evidence that healthcare workers (HCWs) are at higher risk of contracting COVID-19 than the general population, and that Black, Asian, and minority ethnic workers are disproportionately affected [1,2,3]. In April 2020, 6.2% of the National Health Service workforce in England was absent, and between March 2020 and April 2021 the proportion of all working days lost due to COVID-19 ranged from 4 per cent to 30 per cent [4,5]. Data from the UK SIREN study demonstrates that HCWs who are more frequently exposed to those with COVID-19 or who work in higher risk settings have higher infection rates, although exact reasons for this remain unclear [6].

Measures that can be put in place to reduce the spread of COVID-19 in healthcare settings include physical distancing, ventilation and use of personal protective equipment (PPE) such as fluid resistant surgical masks (FRSM) and respiratory protective equipment (RPE; please note the term ‘masks’ will refer to both FRSM and RPE throughout this paper). Masks can prevent transmission by reducing the extent to which COVID-19 is passed on by the wearer (‘source control’) and by protecting the wearer from becoming infected with COVID-19 (‘wearer protection’). NHS HCWs commonly use both FRSM and filtering facepiece (FFP3) respirators based on extant national guidance [7].

There is broad consensus on the recommendation for RPE (i.e., FFP3 masks) when performing aerosol generating procedures (AGPs) [8]. However, outside of these specific procedures, recommendations for RPE use are generally less prescriptive, with international guidance suggesting that the decision to use RPE may be based on several factors including the ventilation in the area, the ability of the patient to wear masks, the vaccination status of the patient, the local prevalence of infection, the availability of RPE, and HCW preference [9,10,11]. Masks can also be used either for a single patient interaction (targeted use) or for a period of time when the healthcare worker is undertaking clinical duties in a specific clinical area (sessional use). NHS England guidance advises that the decision to wear RPE “should be based on clinical risk assessment, e.g., task being undertaken, the presenting symptoms, the infectious state of the patient, risk of acquisition and the availability of treatment for the infectious agent” [12]. International guidance evolved several times throughout the pandemic, initially recommending widespread use of RPE then reserving RPE for AGPs only [9,13,14].

Whatever mask recommendations are in place, for them to be effective compliance with recommended policy must be high. Factors associated with reduced compliance with mask use may include perceived low risk from COVID-19 [15,16], perceived low efficacy of masks for reducing the spread of COVID-19 [17,18], and poor understanding of the rationale for when and why different types of PPE are required [15]. Adverse consequences of wearing masks such as discomfort [19,20] and challenges with communication [21] may also reduce compliance with policy [17]. There may also be potential for self-contamination if masks are not worn correctly, for example by wearers adjusting the mask with contaminated hands, touching their mask or their face, or touching their eyes [22]. It is therefore important to understand HCW experiences of wearing different types of masks, so that any adverse consequences of mask use can be considered alongside any benefits.

### Aims

The current study aimed to address this by exploring HCW experiences and perceptions of wearing different types of masks. Specifically, the aims of the study were to explore participants’: i) understanding of current guidance for wearing different types of masks; ii) perceptions of the efficacy of different types of masks; iii) experiences of wearing different types of masks, in both a targeted and a sessional way; and iv) levels of compliance with wearing different types of masks.

## Method

### Ethical approval

Ethical approval was obtained from the UK Health Security Agency Research Ethics Governance Group (approval no. R&D 496).

### Design

Semi-structured interviews were carried out to understand participants’ perceptions of wearing different types of masks. Interviews took place over Microsoft Teams between 27^th^ May and 26^th^ August 2022 and lasted between 20 minutes and 1 hour and 2 minutes.

### Participants

Participants were healthcare workers (*N* = 12) who were currently working in one of three different hospital trusts in the UK who will be referred to as Trust 1, 2, and 3 for anonymity: Trust 1 (*N* = 1); Trust 2 (*N* = 7); and Trust 3 (*N* = 4). To identify these hospital trusts, an expression of interest for the study was sent to all English sites that were participating in the SIREN study (https://www.gov.uk/guidance/siren-study). The three trusts who participated in the current study expressed interest in taking part and were subsequently sent participant recruitment materials. Participants were then recruited via existing networks of staff in the three participating trusts. Participants were eligible to take part in the study if they were patient-facing healthcare staff in NHS Trusts in England, working in a clinical area with patients with suspected or confirmed respiratory infections, including emergency departments, acute medical wards, and relevant inpatient wards.

### Materials

A semi-structured interview guide was developed to ensure that key points were captured whilst allowing participants to share all relevant experiences. The semi-structured interview guide contained questions relating to participants’: understanding of current guidance for wearing different types of masks; perceptions of the efficacy of different types of masks; levels of compliance with wearing different types of masks; and experiences of wearing different types of masks, for both targeted and sessional use.

### Procedure

Hospitals that had expressed an interest in taking part in the study were contacted and asked to share recruitment information with potential participants (HCWs within each hospital trust who met the inclusion criteria). Potential participants were then asked to contact the research team if they were interested in taking part in an interview. Those who expressed an interest in participating were asked to sign a consent form and a member of the research team then arranged a convenient time for each participant to take part in an interview. Interviews were carried out by behavioural scientists based at the UK Health Security Agency (UKHSA), all of whom were qualified to at least MSc level and had received training in carrying out interviews. Interviews were carried out by both male and female members of the research team and only the researcher and the participant were present during the interview. Researchers did not establish a relationship with participants prior to carrying out the interview nor were participants made aware of any personal characteristics of the interviewer, aside from their place of work and the broad aims of the research. Each interview was recorded and later transcribed for analysis. After taking part in an interview, participants received a debriefing statement and a £25 voucher to thank them for their participation.

### Analysis

Data were analysed using the Framework Approach, a type of thematic analysis which is commonly used in research which has implications for policy and practice [23]. The five steps of Framework Analysis were applied to the data. The first author familiarised themselves with the data, before identifying codes within the data that related to the research questions, in order to create an initial coding framework. Data were then indexed into broad themes, which were discussed with other members of the research team. An analytic framework was then created, before themes were defined and clarified in relation to other themes.

## Results

Six main themes and 13 sub-themes were identified:

1. Perceptions of current mask policy – (i) clarity of guidance; (ii) mandatory vs advisory guidance; and (iii) understanding of why different masks are required
2. Perceived efficacy of FFP3 and FRSM – (i) perceived efficacy for preventing catching and spreading COVID-19; and (ii) perceived efficacy for preventing catching and spreading of other respiratory infections
3. Reasons for different mask preference – (i) comfort associated with wearing each mask type; and (ii) perceived situational risk
4. Preference for targeted vs sessional FFP3 use
5. Difficulties associated with wearing different types of masks – (i) discomfort (ii) difficulty communicating (iii) problems with fit; and (iv) impact on ability to do the job
6. Compliance – (i) overall compliance with mask wearing policies; and (ii) situations which may result in non-compliance.

Additional example quotes to those found in the sections below can be found in Table 1.

**Table 1.**
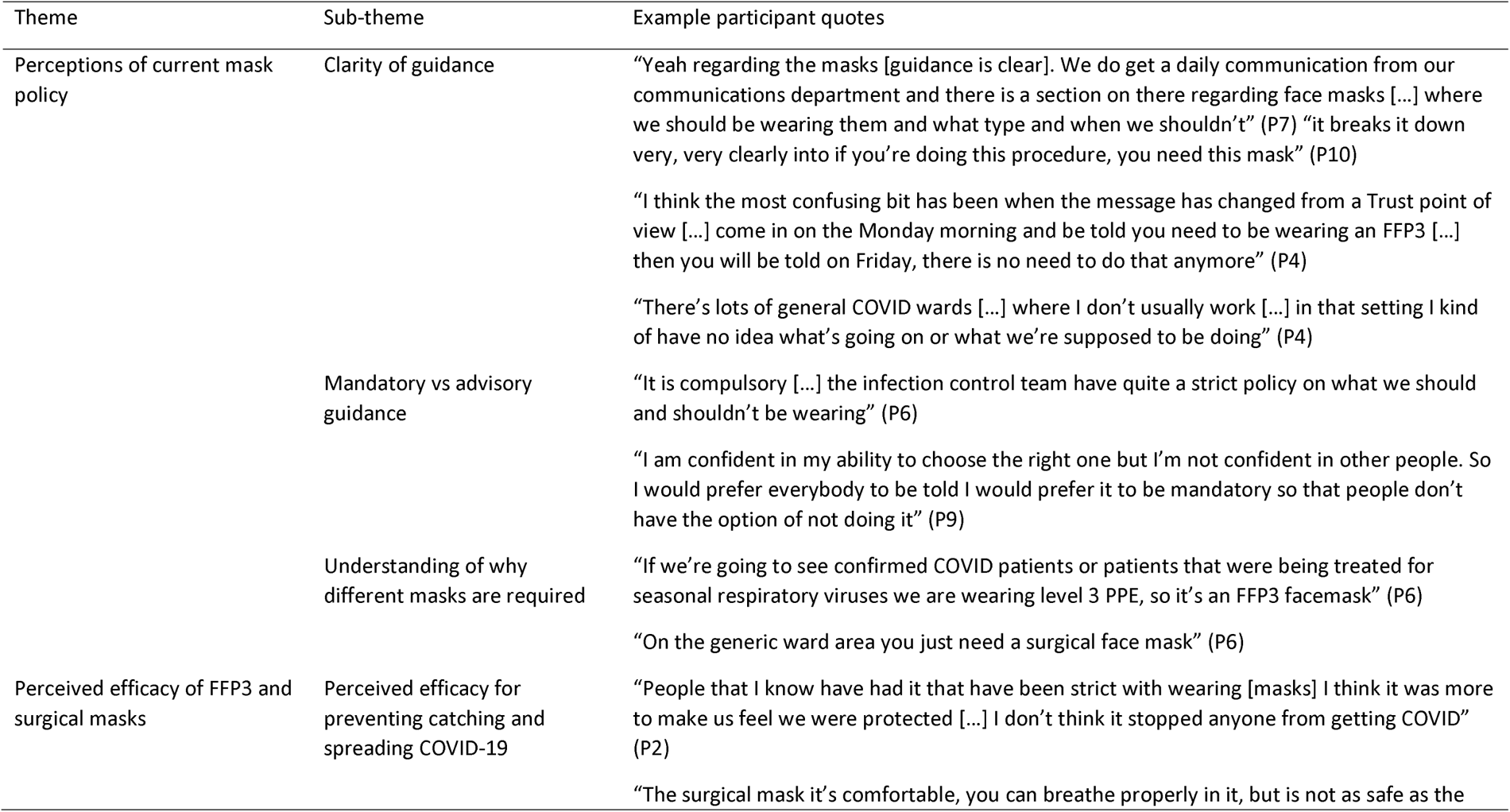
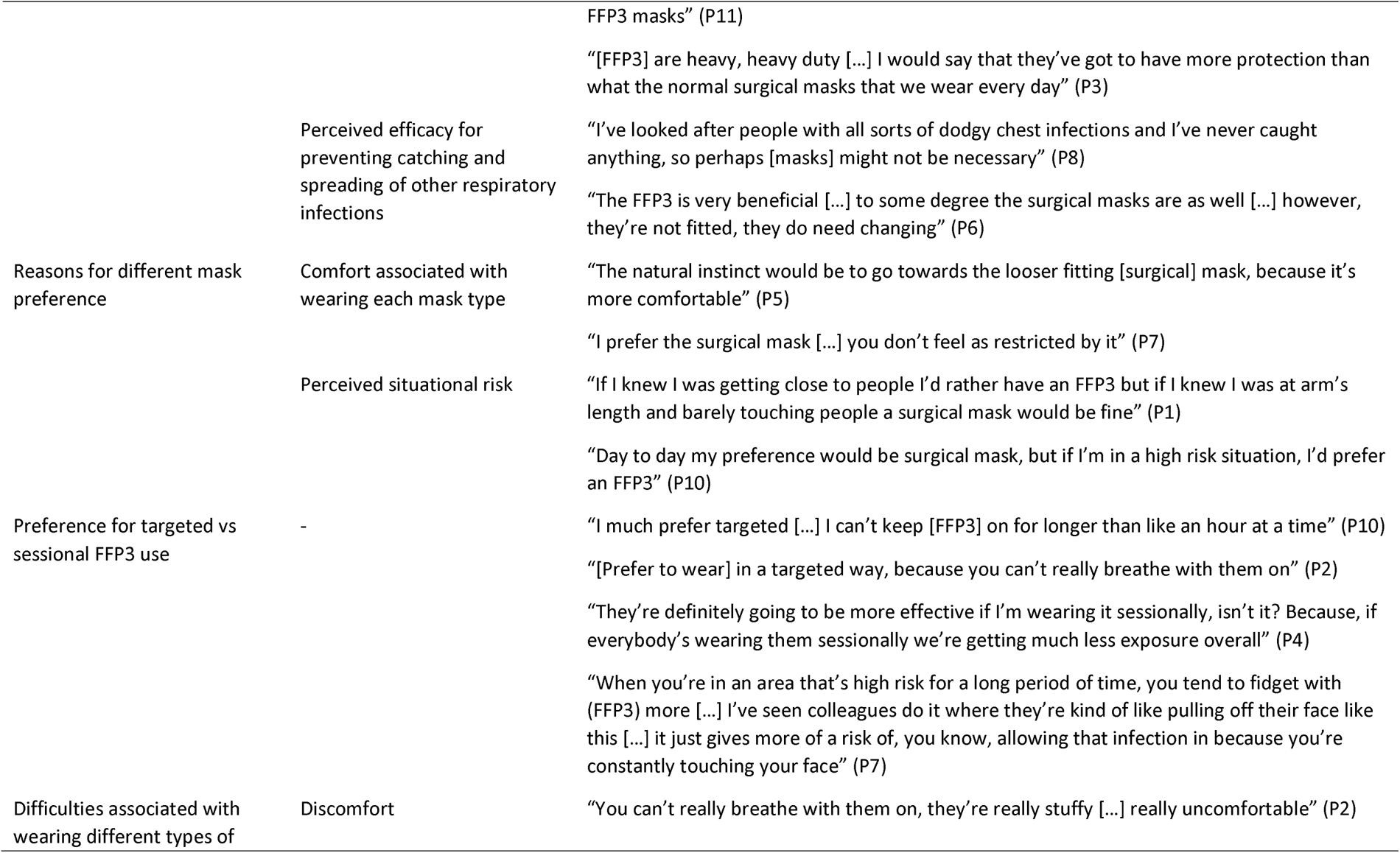
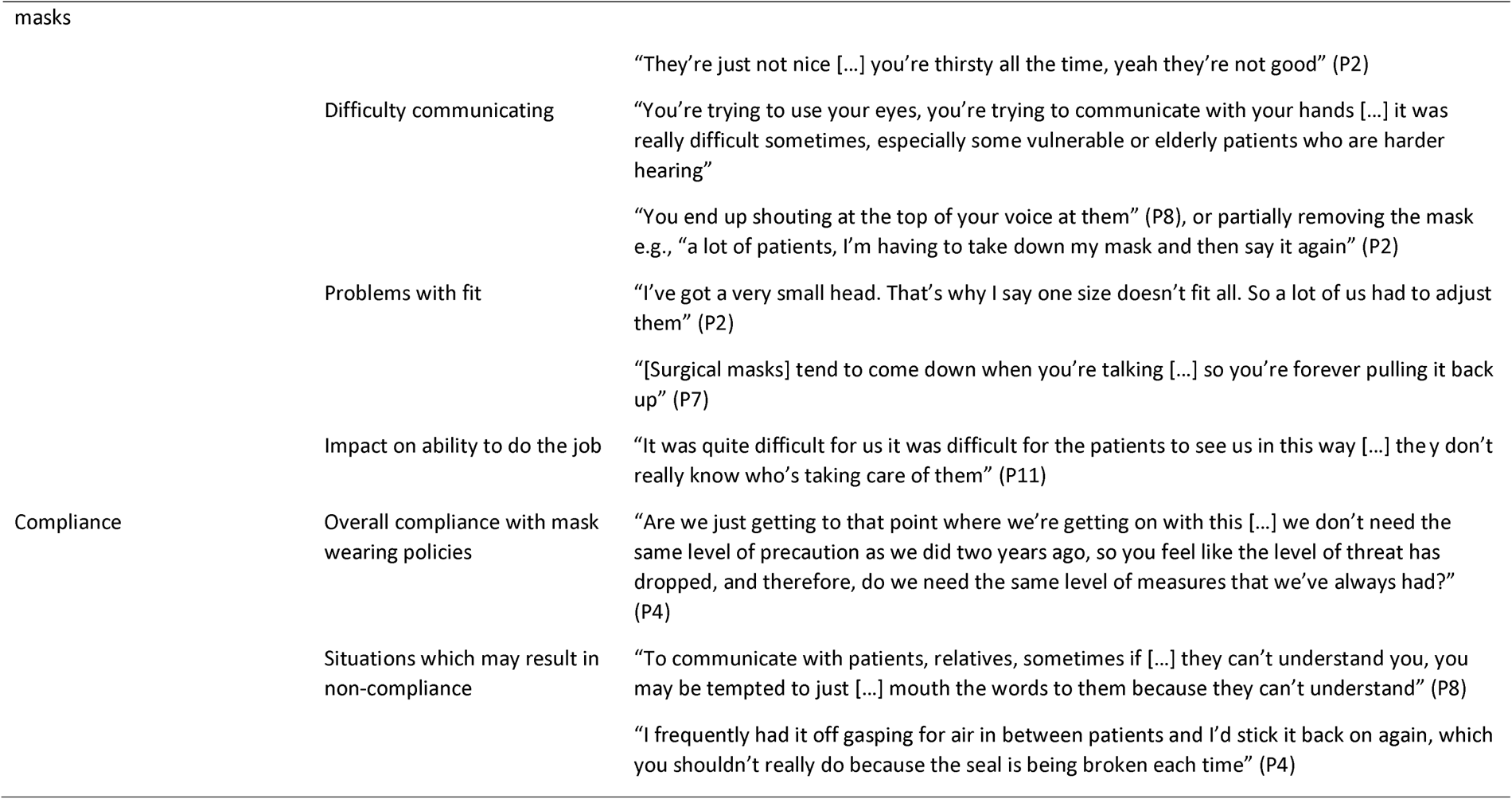
Summary of themes, sub-themes, and example participant quotes.

### Perceptions of current mask policy

Participants were asked how they felt about current policy for wearing different masks within their hospital trusts, including whether the guidance is clear and whether it is mandatory or advisory. Within this theme, three sub-themes emerged: clarity of guidance; perceptions of mandatory vs advisory guidance; and understanding of why different masks are required.

#### Clarity of guidance

Participants expressed different views about whether guidance on mask wearing was clear within their hospitals. Some participants felt that the guidance for mask wearing was clear e.g.“it breaks it down very, very clearly into if you’re doing this procedure, you need this mask” (P10).

However, some participants suggested that the guidance could be clearer. This was particularly the case when guidance changed rapidly e.g., “the Trust has now changed its guidance three times on what to wear […] no, it’s probably not very clear as a Trust” (P9), or when moving to different areas of the hospital in which different policies were in place e.g., “it’s not clear […] I don’t know if it could be made clear because it changes all the time and all the wards change” (P8).

#### Mandatory vs advisory guidance

Most participants stated that guidance is currently mandatory e.g., “It’s mandatory […] you know that we have to wear them in these situations, so mandatory” (P2), though some participants said that guidance is now voluntary e.g., “the guidance at the moment is that if you want to have a mask on that’s down to you” (P5). A few participants were unsure of whether guidance was mandatory or advisory e.g., “to be wearing a mask of some description, I’m sure is mandatory. I just don’t know if it’s mandatory to wear an FFP3 in certain circumstances or not anymore” (P4) or said that this can change depending on where they are working in the hospital e.g., “there might be another setting I go into where it’s absolutely mandatory […] it can change ward to ward depending on the type of patient that’s been treated” (P5).

Participants differed in their views on whether they would prefer guidance to be mandatory or advisory. The main reason participants gave for preferring guidance to be mandatory was that they did not trust others to choose the appropriate type of mask e.g., “I think being told what to wear [is preferable] I think if staff are given an option, then they’re always going to go for the one that […] doesn’t take as much time up or doesn’t take as much energy” (P7). Some suggested that mandatory guidance was preferable, but only if it was based on sensible recommendations e.g., “I’m happy to be told […] but I think that [guidance] needs to be based on sensible recommendations and recommendations that don’t try to prevent the spread of COVID everywhere” (P4). Interestingly, while some felt that mandatory guidance would encourage people to adhere to mask policies, it was also suggested that people may be more likely to follow the guidance if it were advisory e.g., “I’d prefer it to be a kind of guided but make your own decision […] I think people are more likely to follow this guidance […] if that’s the option as opposed to be compulsory” (P10). Other reasons given for preferring advisory guidance were that people felt confident in their ability to choose the correct mask e.g., “I would like to think I’ve had enough experience to know which situations to wear which one in” (P1), and that mandatory guidance is not appropriate in the longer term e.g., “I think more long term, perhaps we should be looking at it more in what the practitioner wants to wear themselves” (P6).

#### Understanding of why different masks are required

All participants stated that they understood why different masks are required in different circumstances e.g., “I am aware of the […] rationale as to why we have different types of PPE” (P6), despite some inaccuracies in understanding of the underlying science e.g., “the surgical masks I understand a bit more general […] but the FFP3’s obviously a bit of a better filter to stop the spores coming through from the infection” (P12). Participants stated that FFP3 masks are required in situations of higher risk, for example when working on a ward with COVID-19 or other respiratory infections e.g., “if we are having any contact with a patient who’s COVID positive then we would be required to wear the FFP3” (P12), or when performing high risk procedures e.g., “yeah [type of mask is] based on risk exposure, so certain medical procedures and certain devices are more aerosol generating and therefore warrant […] a higher value facemask” (P10). In contrast, participants understood that surgical masks are required in situations with lower risk of transmission, such as general clinical areas (as opposed to those with a high risk of COVID) e.g., “so we just wear [surgical masks] now around all our corridors, outpatient and clinical areas” (P7).

### Perceived efficacy of FFP3 and surgical masks

When participants were asked about the perceived efficacy of surgical and FFP3 masks for preventing catching and spreading infections, there were two sub-themes: efficacy for *wearer protection and source control of COVID-19*; and efficacy for *wearer protection and source control of* other respiratory infections.

#### Perceived efficacy for preventing catching and spreading COVID-19

Most participants felt that both surgical and FFP3 masks were effective for preventing them from becoming infected with COVID-19 e.g., “I think they’re effective […] I’ve got colleagues that have not had COVID for the full two years, and now that we’ve stopped wearing the masks, we seem to be catching COVID now” (P3). However, some participants felt that both surgical and FFP3 masks had limited efficacy for preventing them from becoming infected with COVID-19 e.g., “myself and my colleagues […] we’ve constantly worn masks and we’ve still all managed to get COVID” (P7).

Most participants also perceived both surgical and FFP3 masks to be effective, to some extent, for reducing the risk of them transmitting COVID-19 to others e.g., “I’d say it’s just as good at keeping the germs in […] a two-way thing” (P5). However, some participants highlighted that reduced transmission of infection also depended on other types of protective actions, rather than solely mask wearing e.g., “wearing the mask will stop you, you know, from spreading the droplets but if you’ve already coughed in your hand previously and not washed your hand and then touched somebody’s face […] that’s not going to reduce the risk is it” (P7).

The majority of participants perceived FFP3 masks to be more effective than surgical masks for *wearer protection and source control of* COVID-19 e.g., “I don’t think surgical masks have a particularly great impact […] but I do think the FFP3s are definitely worthwhile” (P6). Reasons given for greater perceived efficacy of FFP3 masks included FFP3 masks having a better fit e.g., “surgical masks are a bit more of a transient fit […] they are much tighter fit FFP3s, and I think we know that they’re more effective” (P4) and being thicker e.g., “I’d probably say the FFP3 [is more effective] because it’s a lot thicker” (P2).

#### Perceived efficacy for preventing catching and spreading of other respiratory infections

As with perceived efficacy for wearer protection and source control of COVID-19, most participants perceived the masks to be effective for wearer protection and source control of other respiratory infections e.g., “I think […] they’re quite effective across the board” (P3), “if [the mask is] a barrier to stop COVID it must be doing it’s job, it must be pretty, pretty good at stopping everything else” (P5). However, some were unsure as to the effectiveness of masks for preventing transmission of other respiratory infections e.g., “I wouldn’t know [how effective they are] because I have been at work with colds and things and other people have got colds” (P2). As with COVID-19, most participants perceived FFP3 masks to be more effective than surgical masks for wearer protection and source control of other types of infections e.g., “I think FFP3s are more effective […] I think surgical masks […] it’s not going to be as good as having an FFP3 on” (P4).

### Reasons for different mask preference

There were two main factors that affected participants’ preference for wearing surgical or FFP3 masks: how comfortable each mask type is to wear; and perceived situational risk.

#### Comfort associated with wearing each mask type

Most participants stated that they preferred to wear surgical masks e.g., “If I had to have a preference, it would be just a normal surgical mask” (P3). The main reasons for this were that surgical masks are more comfortable e.g., “surgical masks are nicer, they’re easy to wear, they don’t feel as full on” (P1), and better for wellbeing e.g., “there is the kind of wellbeing and occupational aspect for staff, which has been quite impacting to have to wear an FFP3 for every single patient” (P4).

#### Perceived situational risk

While most participants generally preferred to wear surgical masks, their preference for different mask types varied depending on the perceived risk of the situation e.g., “Surgical more comfortable, FFP3 more protective, just a risk assessment on wherever I am and what I’m doing” (P9). Several participants stated that, though they prefer wearing surgical masks, they would prefer to wear FFP3 masks in high-risk situations e.g., “if I was going to an area which was higher risk […] I would favour an FFP3 in that situation” (P6). This was not the case for all, however, with other participants feeling that FRSM masks provided sufficient protection e.g., “I’d be quite happy with wearing just the normal surgical masks rather than the FFP3 […] I do think they provide us with enough protection without having to wear an FFP3” (P3) or that the risk from COVID-19 has now reduced sufficiently that FFP3 masks should only be used in very specific circumstances e.g., “why would we now be trying to prevent COVID on the scale that we prevented it during the last two years […] I’m very happy to wear FFP3 masks […] where there’s a real need and a patient benefit for it, but […] I’m happy enough to wear the surgical mask” (P4).

### Preference for targeted vs sessional FFP3 use

When asked whether they would rather wear FFP3 masks in a targeted or a sessional manner, the majority of participants stated that they prefer targeted to sessional use. The main reason given for this was the discomfort associated with wearing FFP3 masks for long periods of time e.g., “I do prefer [targeted] I think long term use it’s not comfortable” (P6). It was also suggested that the risk posed by COVID-19 is now not sufficient to require sessional FFP3 use e.g., “the only way I would wear an FFP3 mask again all day long is if we get a new infectious disease, never again for COVID” (P4). However, not all participants preferred to wear FFP3 masks in a targeted way, with the main reason given for preferring sessional use being that they felt safer wearing FFP3 sessionally e.g., “I feel a bit safer with [FFP3] on if I’m honest, so I’d prefer to wear those [sessionally]” (P8). This participant also noted that it could be frustrating to continually have to change the type of mask they were wearing e.g. “I just think can’t be bothered with the faff, just let me wear this, the FFP3, and I’ll just keep it on until I need food or drink” (P8).

Interestingly, although the majority of participants preferred to wear FFP3 in a targeted way, several participants felt that sessional use would be more effective, because it involves wearing the mask for a longer period of time e.g. “sessional would probably work better […] I’d say that the effectiveness probably works better having it on for a longer period of time” (P3), and continually changing the mask risks contamination e.g. “unless you’re meticulous taking your mask off, you’re probably infecting yourself just keep touching your face all the time” (P8). In contrast, some participants felt that targeted use would be more effective. One reason for this was that participants perceived FFP3 to become less effective if worn for long periods of time e.g., “If you use [FFP3] for an extended period of time without changing them […] they won’t last indefinitely” (P1). Another reason that targeted use was perceived to be more effective was that participants felt that people would be more likely to touch FFP3 masks (due to discomfort), creating a risk of infection e.g., “in the sessional use, I’m forever tempted to just pull it down and have 10 seconds of relief. Whereas if I know in targeted use it’s only for 20 minutes, half an hour […] I can cope with it” (P10). One participant specifically highlighted that while sessional use might be more effective, wearing an FFP3 mask for long periods of time would not be in people’s best interest e.g., “As much as I would probably say that the FFP3’s more effective in all settings, and let’s all walk around with them all of the time I also don’t think that’s the best for people” (P12).

### Difficulties associated with wearing different types of masks

When asked about any difficulties associated with wearing surgical and FFP3 masks, participants identified several issues, which fell into four broad categories: discomfort associated with wearing masks; difficulty communicating when wearing a mask; problems with mask fit; and impact of masks on participants’ ability to do their job.

#### Discomfort

Participants described several ways in which they experience discomfort associated with wearing FFP3 masks. These included: difficulty breathing e.g., “you can’t really breathe with them on, they’re really stuffy […] really uncomfortable” (P2); problems with overheating e.g., “when you’re quite active wearing one all day they can make you quite hot” (P12); problems with skin e.g., “I was wearing them for up to six, seven hours sometimes […] it did start to have quite a bad effect on my skin” (P1); feeling claustrophobic e.g., “they do make you feel quite stressed quite […] claustrophobic” (P3); and becoming dehydrated e.g., “I would get home at the end of the day and feel really dehydrated with this horrible dry mouth” (P5).

Participants reported less discomfort associated with surgical masks e.g., “[surgical masks] are fine […] they’re just kind of a normal given everyday for me now” (P10). However, some did report some issues including increased dehydration e.g., “they’re just not nice […] you’re thirsty all the time, yeah they’re not good” (P2), and headaches caused by tight elastic e.g., “the elastic would pull on my ears and I get pain right behind on my skull on both sides” (P5).

#### Difficulty communicating

Participants highlighted that there were communication difficulties associated with wearing both FFP3 masks e.g., “I struggled to hear other people when [they’re] wearing an FFP3, or when they’re wearing an FFP3” (P10), and surgical masks e.g., “you can’t communicate properly with [surgical masks] on” (P8). It was suggested that communication is particularly difficult with those who are vulnerable or otherwise have difficulty communicating e.g., “you’re trying to use your eyes, you’re trying to communicate with your hands […] it was really difficult sometimes, especially some vulnerable or elderly patients who are harder hearing” (P3), and that at times, the need to interact properly with a patient outweighed the risk of infection e.g., “you weigh it up and you think it’s more important that this patient gets an important interaction that he needs than total prevention from COVID” (P4). Participants also spoke about ways in which they tried to overcome difficulties with communication, such as writing things down e.g., “[communication difficulties are] why we got a whiteboard […] but in an emergency that’s like you’re trying to write something down saying can you go and fetch this” (P2), raising their voice e.g., “you end up shouting at the top of your voice at them” (P8), or partially removing the mask e.g., “a lot of patients, I’m having to take down my mask and then say it again” (P2).

#### Problems with fit

Some participants highlighted difficulties associated with mask fit, particularly in relation to FFP3 masks. Difficulties included masks would sometimes fall down e.g., “sometimes the mask would […] drop down on the face, so that was quite difficult” (P11), that FFP3 masks fit some people better than others e.g., “I’ve got a very small head. That’s why I say one size doesn’t fit all. So a lot of us had to adjust them” (P2), and that masks that participants had been fit tested for sometimes ran out, requiring them to then use a mask they had not been fit tested for e.g., “sometimes the masks that we were fit tested on would run out and we’d just use […] one that we were not fit tested on which would put us at risk” (P11). Issues associated with fit of surgical masks were reported less commonly (likely because there is less emphasis on the importance of good fit in relation to surgical masks) but some participants did highlight fit issues associated with surgical masks e.g., “[surgical masks] tend to come down when you’re talking […] so you’re forever pulling it back up” (P7).

#### Impact on ability to do the job

As noted above, participants highlighted that discomfort and communication challenges sometimes impacted their ability to do their job. Participants also highlighted other ways in which wearing an FFP3 mask made doing their job more difficult, including: impact on patient care e.g., “[wearing an FFP3] would probably make me do what I needed to do perhaps in a quicker manner, so that I was in and out” (P6); and difficulty with physically being able to carry out tasks e.g., “I undertake medical photography, so one of the things we have to do is obviously have the ability to focus through the lens […] trying to do that through a visor with an […] FFP3 mask on is very challenging” (P5). While the impact of surgical masks was perceived to be less, some participants did highlight issues that were common to both types of masks, such as glasses fogging up and preventing ability to see e.g., “a lot of my colleagues did struggle because of glasses steaming up” (P7).

### Compliance

When asked about their compliance with mask wearing policies, participants reported two separate aspects associated with compliance: their overall compliance with mask wearing policies; and situations which may result in themselves or others being less compliant.

#### Overall compliance with mask wearing policies

The majority of participants said they were compliant with mask wearing policies, both for FFP3 masks and surgical masks e.g., “I’ve never gone against […] if they say you need to wear one I will wear one” (P5). Reasons for complying included that mask policies are in place for a reason e.g., “I would always wear one […] because I know that on our ward we’re only recommending them when they’re really needed” (P4) and that it is safer to follow the guidance e.g., “we do follow correct PPE guidelines […] to reduce that risk of infection” (P7). Indeed, one participant noted that they sometimes chose to wear a mask even in situations in which they weren’t required, if they felt it was appropriate based on the level of risk e.g., “I chose to actually wear a mask even though I didn’t have to […] because I was in very close proximity of people who were coughing” (P5).

However, there was some suggestion that compliance generally had reduced over the course of the pandemic, with the main reason for this being that the risk has changed e.g., “because people are vaccinated, we are relaxed […] we are more relaxed in wearing FFP3 masks than before” (P11).

#### Situations which may result in non-compliance

Whilst most participants stated that they were compliant with mask wearing policies, they did identify some circumstances under which they would be less likely to comply. A key circumstance under which participants felt compliance would be lower is when the situation was perceived to be lower risk. Factors which contributed to a perception of a situation as low risk included there being no patients around e.g., “we’ve become a little bit more, can I use the word slack […] in an area where there’s no patients” (P4) and people being socially distanced e.g., “I’ve got no one sat next to me this afternoon so I’ve taken my mask off” (P3). Other circumstances under which participants felt compliance may be lower included situations in which communication is challenging e.g., “when we’re having a conversation on the phone, or we feel like someone can’t hear us we pull them down” (P4), and emergency situations in which there may not be time to don or doff the appropriate mask e.g., “it’s sometimes difficult in the moment to don / doff properly when it’s an emergency” (P10). Some also stated that discomfort was a reason for non-compliance e.g., “I was probably cutting corners and pulling it off my face because I just couldn’t cope with it being strapped so tight” (P8).

## Discussion

This study explored HCW experiences and perceptions of wearing different types of masks in healthcare settings. Specifically, the study aimed to explore participants’ understanding of current guidance for wearing different types of masks, perceptions of the efficacy of different types of masks, experiences of wearing different types of masks (in both a targeted and a sessional manner), and levels of compliance with wearing different types of masks. The study focussed on the use of FFP3 masks and FRSM masks.

Findings showed that participants generally had good understanding of when and why different types of masks were required according to extant guidance. Most participants reported that FFP3 masks should be used in situations of higher risk, including when patients have (or are suspected to have) a respiratory virus, and when aerosol generating procedures are required. This is in line with both UK and international guidance that RPE should be worn for higher risk procedures and in higher risk settings and situations [9,10,11]. However, some participants reported that frequent changes to guidance sometimes made it difficult to follow. High levels of understanding are important, as low understanding of mask policy may be associated with reduced compliance [15]. It is therefore important to ensure that guidance on mask policy is communicated clearly, including particularly with updates in the event that guidance has changed.

Participants expressed mixed views as to whether guidance on RPE should be advisory or mandatory. While some would prefer to make their own decisions regarding what type of mask to wear, others felt that they would prefer this guidance to be mandatory. Almost all participants stated that they preferred targeted use of FFP3 masks to sessional use, with the main reason for this being the discomfort associated with wearing FFP3 masks. In particular, participants highlighted that wearing FFP3 masks for long periods of time resulted in discomfort that at times impacted both their ability to do their job and their wellbeing. This is in line with previous research demonstrating that masks have the potential to result in discomfort [20]. Furthermore, participants mentioned that due to this discomfort, they would be more likely to touch and interfere with their FFP3 mask over a prolonged period of time. As such, it could be argued that good adherence to targeted use of RPE may be more effective than poor adherence to sessional use.

The majority of participants perceived FFP3 masks to be more effective than surgical masks for preventing them from catching and spreading COVID-19 and other respiratory viruses. While this is in line with research [24], participants often based these views on their personal experiences, stating that they had not caught COVID-19 whilst wearing FFP3 masks and therefore believed FFP3 masks to be more effective than FRSM’s. This reliance on personal experiences resulted in some participants being uncertain about the role of masks in reducing COVID-19 infection rates, particularly when they, or people they knew, had caught COVID-19. Given the role of perceived efficacy in promoting compliance [17] it is important that HCWs perceive masks to be effective. It may therefore be beneficial to provide HCW with up-to-date research findings relating to the efficacy of different types of masks.

Most participants said they would comply with guidance about what type of mask to wear in different situations. Participants said that a key reason for why they would comply was due to the perceived efficacy of masks for preventing catching and spreading COVID-19 and other respiratory viruses. Participants also mentioned that they would be more likely to comply if they felt the guidance was proportionate to the risk. This is in line with previous research which suggests that perceived efficacy of masks is associated with increased compliance [17]. Interestingly, while most participants reported that they themselves complied with mask guidance, they felt that others were less likely to comply – with some citing this as a reason for preferring mandatory mask wearing policies.

Participants also highlighted some situations or contexts in which they would be less likely to comply with guidance. Key reasons for reduced compliance included difficulties with communicating with patients when wearing masks, particularly with patients who were vulnerable or faced additional communication challenges, and discomfort associated with wearing masks (in particular FFP3 masks). This is in line with previous research which suggests that wearing masks can result in difficulties in communicating with others [21] and that discomfort associated with mask wear is a key factor in reducing compliance with guidance [20]. Participants also reported that they would be less likely to comply with guidance in situations they perceived to be lower risk, and that guidance should be in line with the risk associated with the situation. This is also in line with previous research which demonstrates that perceived risk is a key factor in predicting compliance with wearing a mask [17]. Finally, participants highlighted some factors that made it physically difficult to comply with guidance. For example, participants highlighted that surgical masks tend to fall down regularly and therefore require constant touching in order to readjust. Therefore, while such face touching is against guidance, it is often unavoidable in order to ensure that the mask remains in place.

Whilst this study provides important insights into HCW experiences of wearing different types of masks, there are limitations. First, participants opted into this study, and it is therefore possible that the views of those who opted into the study differ from the views of those who did not. Second, while the HCW in this study worked in a variety of hospital settings, participants were recruited from three hospitals. It would be beneficial for further research to explore HCW experiences and perceptions of mask wear across a wider variety of hospital trusts. Third, while interviews were conducted in late spring / summer of 2022 (during which time the COVID-19 situation in the UK was relatively stable) it is possible that guidance changes across the three months of data collection may have impacted HCW perceptions. Finally, the study relied on self-report descriptions of mask use and adherence to guidance which may bias the findings. As such, future research would benefit from the development of an observational tool for mask use.

## Conclusion and recommendations

Overall, the HCWs in this study reported good understanding of the benefits of FFP3 masks and high levels of compliance with mask policy. However, they highlighted some factors that reduced their compliance with mask policy and impacted their ability to carry out their role. These included wearing masks for longer durations (sessional as opposed to targeted use), guidance being perceived as out of step with risk, and communication challenges and discomfort associated with mask use.

Based on these findings, we recommend: 1) Mask guidance should be communicated clearly, particularly when it has changed; 2) Staff should be familiar with up-to-date research on efficacy of masks, to avoid reliance on personal experiences; 3) Mask guidance should be aligned with risk levels; 4) Masks should be required for as short a duration as possible (based on risk); and 5) Policy should be informed by, or speak to, user experience, concerns, and lived experience of end users.

## Acknowledgements

Holly Carter and Dale Weston are funded by the National Institute for Health Research Health Protection Unit (NIHR HPRU) in Emergency Preparedness and Response (EPR), a partnership between UKHSA, King’s College London and the University of East Anglia and the NIHR HPRU in Behavioural Science and Evaluation in partnership with the University of Bristol. Louise Davidson is also affiliated to the EPR HPRU. Colin Brown is affiliated with the NIHR HPRU in Healthcare Associated Infections and Antimicrobial Resistance, a partnership between UKHSA and Imperial College London. The views expressed are those of the authors and not necessarily those of the NHS, the NIHR, the Department of Health and Social Care or UKHSA. For the purpose of open access, the author has applied a Creative Commons Attribution (CC BY 4.0) licence to any Author Accepted Manuscript version arising.

## Data availability

Data are available upon reasonable request.

